# Modeling the microbial contribution to human Energy Balance using the Digestion, Absorption, and Microbial Metabolism (DAMM) model

**DOI:** 10.1101/2025.01.10.25320296

**Authors:** Taylor L. Davis, Blake Dirks, Elvis A. Carnero, Karen D. Corbin, Steven R. Smith, Andrew Marcus, Rosa Krajmalnik-Brown, Bruce E Rittmann

## Abstract

Colonic microorganisms have been linked to human health and disease, specifically metabolic disease states such as obesity, but causal relationships remain to be established. Previous work demonstrated that interactions between the host’s diet and intestinal microbiome were associated with human energy balance by affecting the human’s energy absorption, quantified by metabolizable energy. We developed the Digestion, Absorption and Microbial Metabolism (DAMM) model, which explicitly accounts for the energy contributions of the colonic microbial community by: 1) breaking down the diet composition into the gross energy of the individual macronutrients, 2) calculating direct absorption in the upper gastrointestinal tract, 3) using microbial stoichiometry to estimate the consumption of the remaining unabsorbed nutrients by microbes in the large intestine, 4) quantifying predicted production of microbial products (short-chain fatty acids (SCFA) and methane) in the colon, and 5) estimating absorption from the colonic tract to the host. When used to predict the results from a clinical study that compared two distinctly different diets, the DAMM model captured the directionality and magnitude of change in measured metabolizable chemical oxygen demand (which can be converted to metabolizable energy), improved on the accuracy of predictions compared to the Atwater factors by reducing systematic bias on one of the diets, and estimated substrate availability within the colon and rate of production of microbially derived short-chain fatty acids. Measured methane concentrations, combined with findings from the DAMM model, support the hypothesis that methanogens accumulated in mucosal biofilms in participants harboring methanogens. Model outputs also support that colonic transit time directly influenced SCFA absorption rates. The DAMM model now can be linked to existing human models that predict changes in body energy stores to extend our understanding of how microbial metabolic processes affect macronutrient absorption and metabolizable energy.

## Introduction

The microorganisms that live within the human colon metabolize undigested food and contribute to the energy available to the human host from ingested food (1). Gut microbes use hydrolases that human cells do not possess to break down complex food polymers, and then bacteria ferment the monomers to simpler products that can be easily absorbed in the colon by the human host or can be excreted in feces (1,2). The literature highlights numerous connections between colonic microorganisms and human health and disease, including metabolic diseases such as obesity and diabetes (3–6), autism (7), and even psychological/emotional states (8). In humans, the exact mechanisms by which microbes influence the development of or protection from obesity are not established, though qualitative relationships between specific microbes and obesity are beginning to be investigated (9).

A fundamental concept within the study of human energy balance is metabolizable energy (E_m_), which is defined as the energy from food that is available for the human’s metabolic processes (10). However, in environmental microbiology, the standard measurement for microbial substrates is chemical oxygen demand (COD) (11). COD is the measured mass of oxygen required to completely oxidize organic carbon and represents more generally electron equivalents (11); the use of electron equivalents is important in environmental microbiology because it can be used with microorganisms which, unlike human cellular respiration, are not limited to oxygen as an electron acceptor. In our prior work, we developed a set of regression equations to convert between E_m_ and metabolizable chemical oxygen demand (COD_m_) (12).

In the late 19^th^ century, Atwater and co-workers developed an empirical relationship to calculate E_m_ as a fraction of gross energy (E_g_), defined as the total combustible energy within food items (13–15). The Atwater system uses a single factor to convert the mass of food (grams) directly to the E_m_ (kcal or kJ) for each macronutrient, i.e., proteins, carbohydrates, and fats; the factors are 4 kcal g^-1^ for proteins, 4 kcal g^-1^ for carbohydrates, and 9 kcal g^-1^ for fats (13,15). Due to their simplicity and reasonable accuracy, the Atwater factors remain the most commonly used method to calculate E_m_ (14) and there have been efforts to improve the factors by accounting for fiber fermentation (16).

The three Atwater factors were developed empirically by subtracting energy of combustible gas, fecal, and urinary excretions from measurements of the heat of combustion of total ingested proteins, carbohydrates, and fats (14). The method is unable to account for interpersonal variations, including differences in diet that affect the colonic microbiome. In particular, the Atwater method lumps many metabolic factors into one parameter. A key “lumped-in” factor is the organic acids produced by the colonic microbiome and then absorbed and metabolized by the human host. The Atwater factors *de facto* assume that the microbial contribution is consistent for all persons, even though the microbial community is known to vary with sex, age, race, geographical region, genetics, the health of the host, and, especially, diet (17–22). Several studies found that the Atwater E_m_ calculation systematically overestimates or underestimates E_m_ for various food items by not accounting for differences that affect the gut microbiome (10,13,23–26). In addition, the Atwater approach does not separate the absorption of dietary substrates in the upper gastrointestinal tract from the lower gastrointestinal tract; thus, it does not explicitly include the organic acids produced by the metabolism of the colonic microbiome.

A model that considers the microbial contribution to host metabolizable energy – by tracking food-related energy throughout the digestion and metabolic processes – can be used to understand and quantify the interactions between the gut microbiome and the human body. Here, we describe our Digestion, Absorption, and Microbial Metabolism (DAMM) mathematical model, which tracks the progression of ingested macronutrients as they advance through the host’s digestive tract until they are subsequently absorbed and metabolized by organs and tissues or excreted. Most importantly, the integrated mathematical model explicitly accounts for the contribution to COD_m_ – which can be directly converted to E_m_ (12) – by the colonic microbial community. Using the DAMM mathematical model, we quantify how diet affects the products of microbial metabolism and the total metabolizable COD_m_ (and, by conversion, kcal (12)) absorbed by the host. This mathematical model provides novel insights into the role of microbial metabolisms in host digestion while improving accuracy of metabolizable COD_m_ predictions found in alternate prediction methods (15).

## Methods

### DAMM model concept and structure

The DAMM model was developed using bioreactor principles, with the model’s coefficient values, definitions, and sources detailed in the **Supplemental Material**. A flow chart illustrating the primary processes considered in the DAMM mathematical model is shown in **Figure 1**. The principal function of the model is to predict the metabolizable chemical oxygen demand (COD_m_) that is absorbed in the Upper Gastrointestinal Tract (UGI) and Lower Gastrointestinal Tract (LGI).

**Figure 1:**
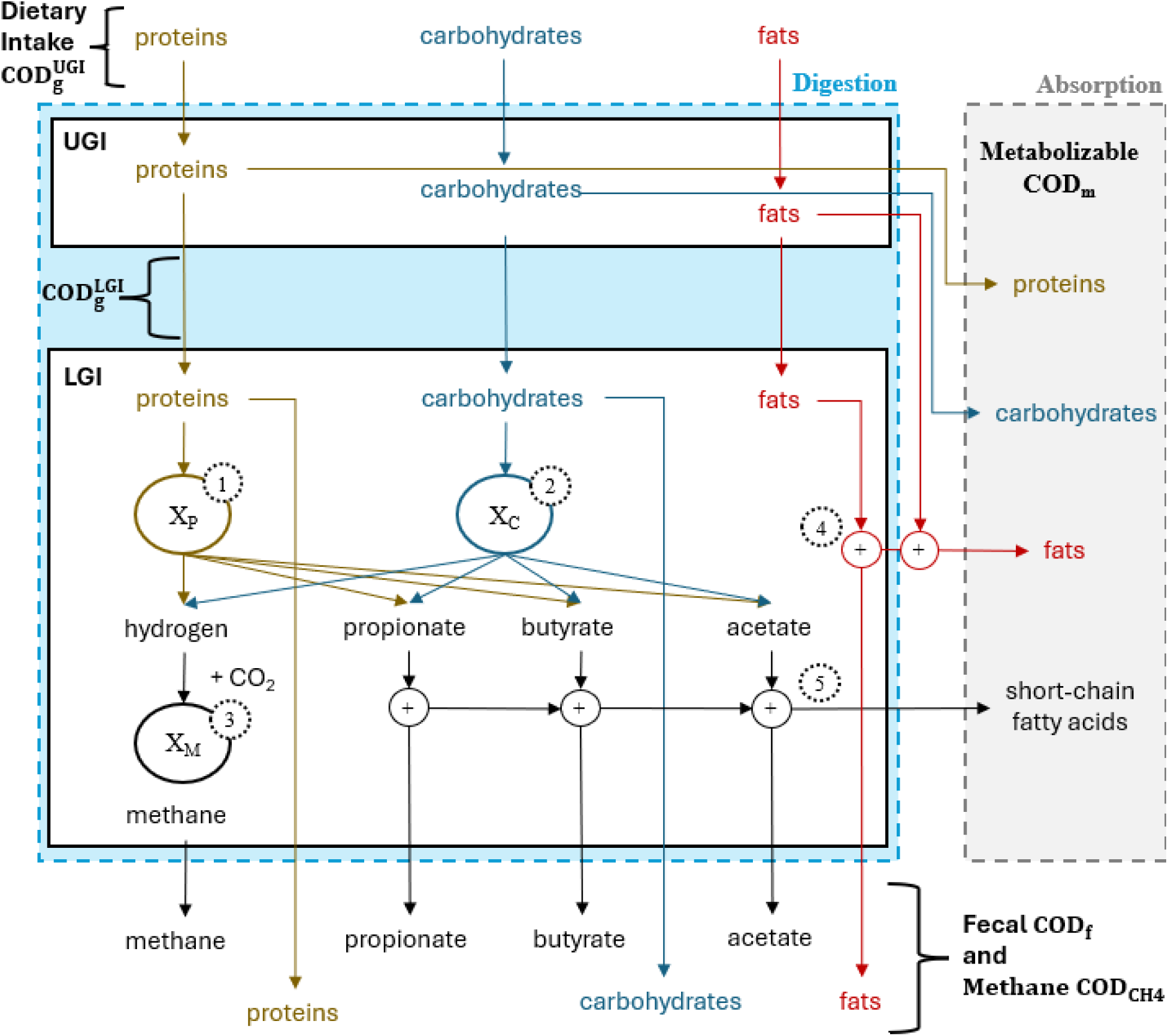
Flow chart of the DAMM mathematical model. The diagram tracks proteins, carbohydrates, and fats as they travel through the digestive tract and then are output either as feces or gas excretions or absorbed by the human body. Processes labeled with numbers in dotted circles are 1) hydrolysis and fermentation of proteins, 2) hydrolysis and fermentation of carbohydrates, 3) methanogenesis, 4) absorption of fats, and 5) absorption of short-chain fatty acids (SCFAs). The blue dotted outline defines the control volume for the digestive tract, while grey is the rest of the human body. UGI and LGI are the upper and lower gastrointestinal compartments, respectively. COD is chemical oxygen demand, while subscripts refer to the following: g – gross, m – metabolizable, f – fecal, and CH_4_ – methane.

While the DAMM mathematical model is founded on a previously published version of the digestion model (27), it has major improvements. Specifically, we included a more comprehensive description of the consumed macronutrients, refined the stoichiometry and kinetics of the colonic microbial community, incorporated methanogenesis as a potential reaction, and considered absorption of short-chain fatty acids (SCFAs) by colonic epithelial cells. These changes are illustrated in **Figure 1**, along with the output streams to the labeled Absorption partition. Here, we discuss the model’s diet inputs, other inputs, processes, and outputs.

### Dietary intake as chemical oxygen demand converted from grams of ingested macronutrients

The DAMM model’s diet input is the daily mass of macronutrients (proteins, carbohydrates, and fats) consumed by the participant in gCOD d^-1^ and labeled as COD_g_^UGI^ (top of Figure 1). We used Equation 1 to convert the mass of each food item (amt_food_ [g_food wet weight_ d^-1^]) to the daily gross COD of each nutrient (COD_g_^nutrient^ [gCOD d^-1^]) for the input.

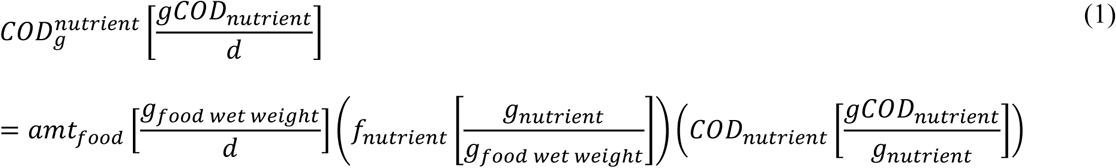

The mass fraction of each macronutrient (amino acids, lipids, sugars, starch, and fiber) in the food item (f_nutrient_ [g_nutrient_ g_food wet weight_^-1^]) was estimated using the United States Department of Agriculture (USDA) Food Data Central Database (28); the database contains the gram amount of individual amino acids, lipids, sugars, starch, and fiber quantities per 100 grams for thousands of food items and was used by the menu creation software when designing the two clinical trial diets (29). The daily ingested mass of each macronutrient in a specific food item was determined by multiplying the daily intake of that food item by the fraction of the macronutrient it contains. This was then converted to COD_g_^nutrient^ by multiplying the value by the specific chemical oxygen demand of the nutrient (COD_nutrient_ [gCOD g^-1^]). The values for COD_nutrient_ were calculated from the chemical formula of each macronutrient using established procedures described in our prior publications (11,12).

Using Equation 1 for all food items ingested per day resulted in a comprehensive breakdown of the composition of the diet, since the ingested COD of each macronutrient was known. This level of detail for a diet is a substantial advancement from the prior versions of our digestion model (22,27), which aggregated the diet input into these categories: protein, fat, non-fiber carbohydrate, and fiber carbohydrate.

### Measured colonic transit time and initial methanogen biomass

We input into the DAMM model two other state variables: colonic transit time (CTT [d]) and an estimate of the initial methanogen biomass within the colon (X_M_^0^ [gCOD]). During our clinical trial (22), the details of which are listed in a later section, the CTT was measured using a radio-transmitting capsule (SmartPill™; Medtronic, Minneapolis, MN) that recorded internal temperature, pressure, and pH (22,29,30). The data were analyzed to determine CTT for each participant on each diet; CTT was identified as a parameter with a complex and highly variable relationship to diet for individuals. Within the model, CTT is incorporated into the equations for microbial production rates and SCFA absorption.

Methanogenesis does not occur in all individuals and requires the presence of methanogens within the colonic microbial community (31,32). To model methane (CH_4_) production, the DAMM model requires an initial estimate of the methanogen biomass COD (X_M_^0^ [gCOD]). We used data generated with quantitative polymerase chain reaction (qPCR) targeting the gene for the methyl coenzyme M reductase (*mcrA*) to measure the gene copy number of methanogens for each participant’s fecal samples (31). From the *mcrA* gene copy number, we estimated an initial methanogen biomass using the following equation:

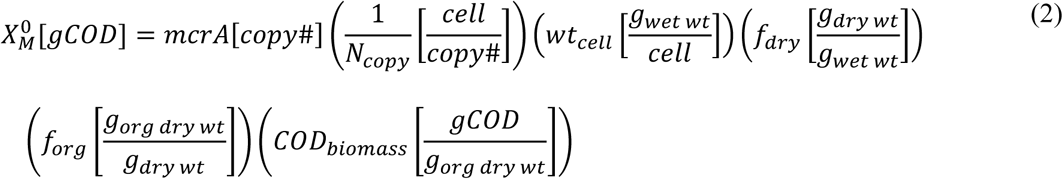

*Methanobrevibacter smithii* represented the methanogen population, based on its being prominent in the intestines (32,33) and detected in our study (31). We estimated the organic dry weight per cell of *M. smithii* as 2×10^-13^ g cell^-1^ using “rod” dimensions (length = 1.5 μm, diameter = 1 μm, density = 1 g cm^-3^ (34)) and assumed 20% of the cell is organic dry mass (11). The methanogen biomass was represented using the formula C_5_H_7_O_2_N (11), making the specific biomass COD_biomass_ [gCOD g^-1^] 1.42 gCOD g^-1^ for the methanogens. Parameters used in the calculations are summarized in **Table 1**, and calculated values by participant are in **Supplemental Material Table 1**.

**Table 1:** Methanogen-specific parameters. List of methanogen-specific parameters utilized in Equation 2 to calculate initial methanogen biomass (X_M_^0^) from the methyl coenzyme M reductase (*mcrA*) copy number measurements. Each parameter has its value with units, a description of the parameter’s purpose, and the source for the value.

| Variable | Value | Units | Description | Source |
| --- | --- | --- | --- | --- |
| $N_{copy}$ | 1 | copy# cell<sup>-1</sup> | Gene copy # per cell for methanogens | (35) |
| $wt_{cell}$ | $1.2 \times 10^{-12}$ | g<sub>wet wt</sub> cell<sup>-1</sup> | Cell weight V*ρ, assume a cylinder for volume: h = 1.5 μm, d = 1 μm then V = $1.2 \times 10^{-12}$ cm<sup>3</sup>/cell and ρ = 1 g/cm<sup>3</sup> | (34) |
| $f_{dry}$ | 0.2 | g<sub>dry wt</sub> g<sub>wet wt</sub><sup>-1</sup> | Dry weight per gram wet weight | (11) |
| $COD_{biomass}$ | 1.42 | gCOD<sub>meth</sub> g<sub>org dry wt</sub><sup>-1</sup> | Specific biomass COD, calculated with cell chemical formula as C<sub>5</sub>H<sub>7</sub>O<sub>2</sub>N | (11,27) |
| $f_{org}$ | 0.9 | g<sub>org dry wt</sub> g<sub>dry wt</sub><sup>-1</sup> | Organic fraction of dry weight | (11) |

### Absorption of proteins, carbohydrates, and fats in the upper gastrointestinal tract

The model’s UGI compartment estimated the absorption of proteins, fats, and carbohydrates in the small intestine before entering the colon (UGI portion of Figure 1). The UGI model was adapted from a model used to simulate the human digestive tract post-colonic-resection surgery (27). Because the participants in the parent clinical trial had an intact GI tract, we removed the effect of the loss of a portion of the small intestine, which gave the rate of COD absorption in the UGI (COD_m_ ^UGI^ [gCOD d^-1^]) as:

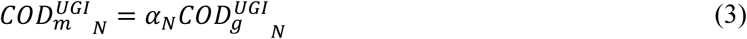

which can be applied for any macronutrient *N* (i.e., fiber, starch, or any of the sugars, amino acids, or saturated and unsaturated lipids). The variable *α_N_* represents the absorption fraction of nutrient *N*, and the variable COD_g_^UGI^_N_ is the total ingested amount of the nutrient, in gCOD d^-1^. Specific UGI-digestibility fractions for each nutrient were taken from the literature (27,36–38); **Supplemental Material Table 2** presents the UGI constants.

**Table 2:** Kinetic parameters. List of kinetic parameters for methanogenesis, carbohydrate, and protein hydrolysis and fermentation utilized in Equations 3-12 to calculate the amounts of microbially produced products within the LGI compartment of the DAMM model.

| Variable | Value | Units | Description | Source |
| --- | --- | --- | --- | --- |
| $k_{F\ abs}$ | 0.48 | $d^{-1}$ | First-order absorption coefficient for fat in the LGI | (27) |
| $k_{C\ ferm}$ | 10 | $gCOD^{-1}\ d^{-1}$ | Second-order kinetic coefficient for fermentation of simple sugars like glucose or fructose | (11) |
| $k_{C\ hyd}$ | 1 | $gCOD^{-1}\ d^{-1}$ | Second-order kinetic coefficient for hydrolysis of complex carbohydrates like fiber or resistant starch | (11) |
| $k_{P\ hyd}$ | 5 | $gCOD^{-1}\ d^{-1}$ | Second-order kinetic coefficient for hydrolysis of proteins into amino acids | (11) |
| $k_M$ | 0.5 | $gCOD^{-1}\ d^{-1}$ | Second-order kinetic coefficient for methanogenesis | (11) |
| $K_{scfa\ abs}$ | 0.02 | $d$ | Half-saturation constant for absorption of SCFAs in the LGI | (30) |

Equations 4 and 5 give the steady-state nutrient-COD balance for the UGI:

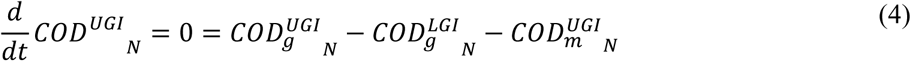

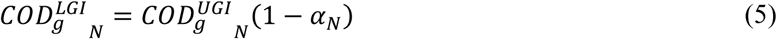

COD_g_^UGI^_N_ is the total ingested nutrient COD, COD_g_^LGI^ is the nutrient COD going into the colon, and COD ^UGI^ is the nutrient COD absorbed within the small intestine. The model operates on the timescale of a 24-hour day, while the orocecal transit time (time from mouth to the beginning of the colon) is approximately 5 hours (39,40); thus, we assumed no storage of nutrients within the UGI (*d/dt COD^UGI^_N_* = 0), which means that the nutrient was either absorbed in the UGI or entered the colon. The output from the UGI compartment was the input for the LGI compartment.

### Lower gastrointestinal tract mass balance

The LGI compartment encompasses the physiological processes of the colon and the colon’s microbiome: microbial fermentation reactions of proteins (Figure 1: process 1) and carbohydrates (process 2) to SCFA products, methanogenesis (process 3), absorption of fat within the colon (process 4), and absorption of SCFAs within the colon (process 5). We represented the colon as a single continuously stirred reactor (CSTR) with a solids’ retention time (SRT) equivalent to the measured CTT (11). We also tested a plug-flow reactor, but found no statistical difference between the CSTR and plug-flow reactor results, consistent with the findings of Moorthy and colleagues (41,42). Thus, we used the CSTR configuration.

A CSTR mass balance equation was used to solve for the effluent mass leaving the LGI compartment:

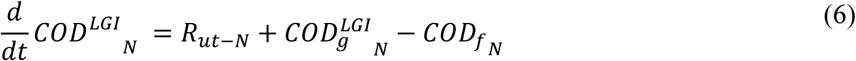

The variable R_ut-N_ [gCOD d^-1^] is the total utilization rate per day for the macronutrient *N*; we included the five processes described in Figure 1 in this term. The last two terms express the influent (*COD_g_^LGI^* [gCOD d^-1^]) and effluent flow (*COD_f N_* [gCOD d^-1^]), respectively. Due to the perfectly mixed assumption of the CSTR configuration, the LGI effluent concentration is equivalent to the concentration within the reactor; thus, the LGI effluent flow is equivalent to the concentration within the reactor divided by the solids’ retention time.

### Microbial hydrolysis and fermentation of proteins and carbohydrates

The LGI model considers microbial hydrolysis and fermentation of proteins and carbohydrates. The microbial utilization of protein is labeled process 1 in Figure 1. We considered protein metabolism in the LGI to occur in two stages: hydrolysis, which is the rate-limiting step, followed by fermentation that produces H_2_, propionate, butyrate, and acetate. A mixed second-order rate described the rate of protein hydrolysis (*R_P hyd_* [gCOD_N_ d^-1^]):

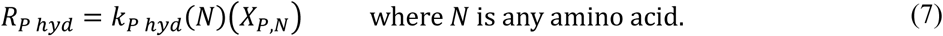

The variables *k_p hyd_* [gCOD ^-1^ d^-1^], *N* [gCOD_N_], and *X_P, N_* [gCOD_X_] are, respectively, the second-order rate constant for protein hydrolysis, the amount of COD for amino acid *N* in the colon, and the current amount of protein-degrading biomass COD using substrate *N* in the colon. Hydrolysis constants are listed in **Table 2**.

For utilizations of fiber and resistant starch (complex carbohydrate), we assumed the same two-step process, but the products were limited to the three main SCFAs: propionate, butyrate, and acetate. However, simple carbohydrates do not require hydrolysis; thus, we defined a separate rate equation for simple carbohydrate fermentation. Carbohydrate hydrolysis and fermentation are collectively labeled as process 2 in Figure 1. We used the mixed second-order rate equation to describe the hydrolysis of complex carbohydrates and the fermentation of simple carbohydrates.

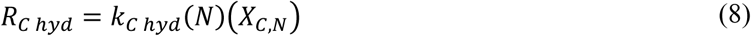

where *N* is a fiber or starch

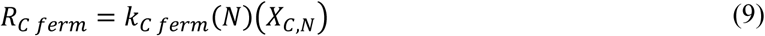

where *N* is a sugar.

The variables *k_C hyd_* [gCOD_X_ ^-1^ d^-1^], *k_C ferm_* [gCOD_X_ ^-1^ d^-1^], and *X_C, N_* [gCOD_N_] are, respectively, the second-order complex carbohydrate hydrolysis rate constant, the second-order simple carbohydrate fermentation rate constant, and the current amount of carbohydrate-degrading biomass COD within the colon using carbohydrate *N* as a substrate. In Equation 8, *N* [gCOD_N_] is either fiber or starch COD in the colon, and in Equation 9, *N* [gCOD_N_] is simple-carbohydrate COD in the colon. Hydrolysis and fermentation constants are listed in **Table 2**.

We used an energy-and-electron-balancing method (29) to develop the stoichiometry matrix for carbohydrate and protein fermentation. The product ratios for each amino acid nutrient were taken from Ramsay et al. (44). Product ratios for carbohydrate fermentation were the same as in Marcus et al. (27). To include biomass synthesis in the stoichiometry, we assumed the fraction of donor electrons utilized for cell synthesis was equal to 0.2 (i.e., f_s_ = 0.2 e^-^ to synthesis/e^-^ from the donor), a value estimated from known values of anaerobic fermenters (11). In the reaction stoichiometry, any type of microbial biomass was represented using the empirical chemical formula C_5_H_7_O_2_N. The stoichiometry matrix in gCOD is in **Supplemental Material Table 3**.

**Table 3:**
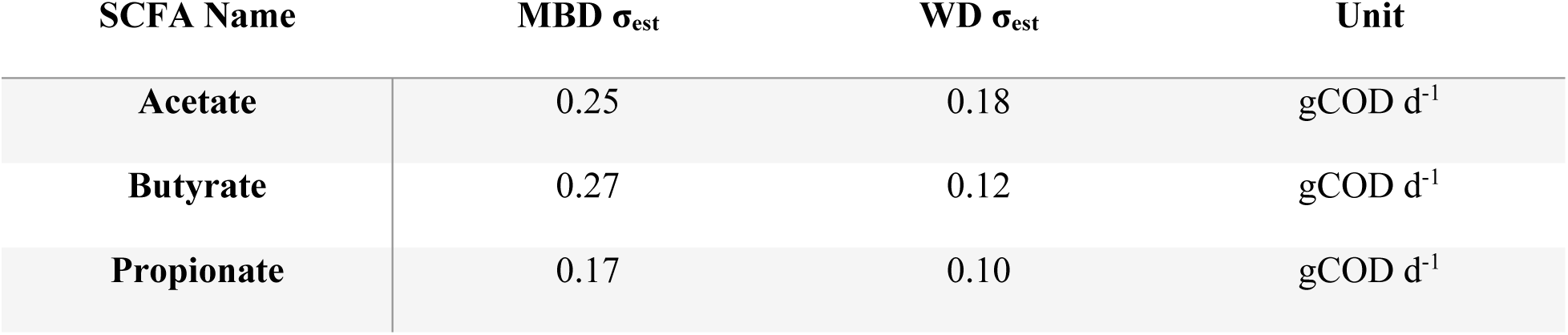
Standard error of estimate (σ_est_) for DAMM predictions of fecal SCFA COD. The error of estimate for SCFA predictions for the identity line i.e., that prediction should equal measurements.

### Methane production if methanogens are present in the lower gastrointestinal tract

Though methanogens are low-abundance species within the gut microbiome, they are one of three identified microbial groups that consume H_2_, which strongly influence microbial interactions (1,20,45). All known methanogen species in the human gut oxidize H_2_ and reduce CO_2_ to produce CH_4_, which is then excreted by the human host (Figure 1: process 3). Within the clinical trial, we measured the total CH_4_ released by the human over the course of 23 hours in a whole-room indirect calorimeter (46). For CH_4_ calculation, the equation for methanogenesis stoichiometry was developed using the same method detailed in the carbohydrate and protein hydrolysis and fermentation sections assuming hydrogen as the electron donor and acetate as the carbon source (47); the complete stoichiometry table is in **Supplemental Material Table 3** and step by step calculations for the stoichiometry and yield were included in the **Supplemental Materials** section titled **Methanogen Yield Calculations**. Similarly, we used the same mixed second-order rate as for the other microbial processes.

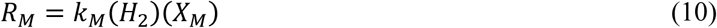

The variables *k*_*M*_ [gCOD^-1^ d^-1^], *H*_2_ [gCOD], and *X*_*M*_ [gCOD] represent, respectively, the methanogenesis second-order rate constant (**Table 2**), the COD equivalent of H2 produced by microbes present in the colon, and the COD of methanogen biomass.

### Absorption of short-chain fatty acids and fats in the lower gastrointestinal tract

For carbohydrates and proteins, a human’s direct absorption (COD_m_ ^UGI^), i.e., not requiring hydrolysis and fermentation by the gut microorganisms, occurs in the UGI tract (Figure 1). Direct absorption of fats (Figure 1) is not restricted to the UGI. We define the rate of fat absorption, R_F abs_ [gCOD_N_ d^-1^], in the LGI using first-order kinetics:

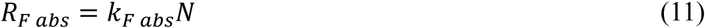

In Equation 11, *N* [gCOD_N_] is the mass of a specific fat, and *k_F abs_* [d^-1^] is the first-order rate constant. We calculated the absorption of microbially produced SCFAs within the LGI (Figure 1: process 5). In the prior version of the digestion model (27), we assumed the absorption of SCFAs within the colon is a constant proportion of 95%, but we also showed that the amount of SCFAs absorbed from the colon was affected by the CTT (48). We computed the fraction of SCFA absorption using a saturation relationship based on CTT:

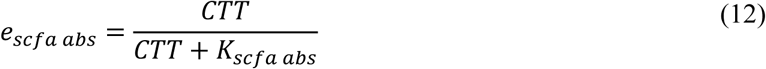

which is illustrated in **Supplemental Material** Figure 1. In Equation 12, *e_scfa abs_* is the mass of absorbed SCFAs normalized to the total mass of produced SCFAs, CTT [d] is the colonic transit time, and *K_scfa abs_* (**Table 2**) is the CTT when 50% of SCFAs are absorbed.

### DAMM model validation – clinical trial data and statistics

We evaluated the DAMM model’s capabilities by comparing its outputs to data from a clinical trial (NCT02939703) designed to quantify the impact of the gut microbiome on human energy balance (22,29). Within the clinical study, 17 healthy men and women consumed two diets in a randomized crossover design. Each diet was consumed at home for 11 days and then for 11 days while the participant was domiciled in a metabolic ward. A ≥ 14-day washout period occurred between the two diet periods. The microbiome enhancer diet (MBD) was a “whole-foods” diet high in dietary fiber and resistant starch but minimally processed foods. The MBD was designed explicitly to maximize the delivery of fermentable dietary substrates to the colon. In contrast, the western diet (WD) was high in processed foods and low in dietary fiber and resistant starch. Both diets were designed to provide energy (kcals) to match the participant’s measured energy expenditure over 6 days in a whole-room calorimeter to achieve energy balance on both diets. Meals were designed by a Registered Dietitian using research-grade software (ProNutra Version 3.5, Viocare, Inc, Princeton, NJ) and prepared in a metabolic kitchen to precisely deliver the nutrients as designed. Adherence was >99% on both diets, and any food not eaten was weighed so that the actual nutritional intake could be calculated. Fecal samples were collected for the 6 days when the participants resided within the whole-room calorimeter.

We used several statistical metrics to evaluate the DAMM model’s performance. For direct comparison between measured and predicted values, we evaluated the accuracy of the predictions using the coefficient of determination (R²) and the standard error of the estimate (σ_est_) relative to the identity line (X = Y). When comparing the metabolizable COD_m_ predictions provided by the DAMM model with the Atwater constants, we used a Bland-Altman plot to visualize the agreement between the predictions and measurements. We looked at the systemic bias by doing a one-sample t-test against the zero difference between methods and calculating Kendall’s coefficient of concordance (τ) for proportional bias. All statistics were computed using the SciPytools package in Python (49).

## Results & Discussion

### Model predicts microbial contribution to metabolizable COD from upper and lower gastrointestinal tracts

The DAMM model tracks the flow of COD from macronutrients as they move through the upper and lower gastrointestinal tracts. Figure 2A presents the average daily distribution of COD – with pie plots showing the breakdown into protein, carbohydrate, fat, and SCFAs – across the inlet and outlet streams depicted in Figure 1 for both diets. The dietary intake, i.e., gross COD for the UGI inlet (COD_g_^UGI^), had an average value of 630 ± 105 gCOD d^-1^. The DAMM predicted that 102 ± 21 gCOD d^-1^ (16.2 ± 2.1% of dietary intake) continued into the colon, while only 26 ± 8 gCOD d^-1^ (4.2 ± 0.9% of dietary intake) was excreted as feces or gas.

**Figure 2:**
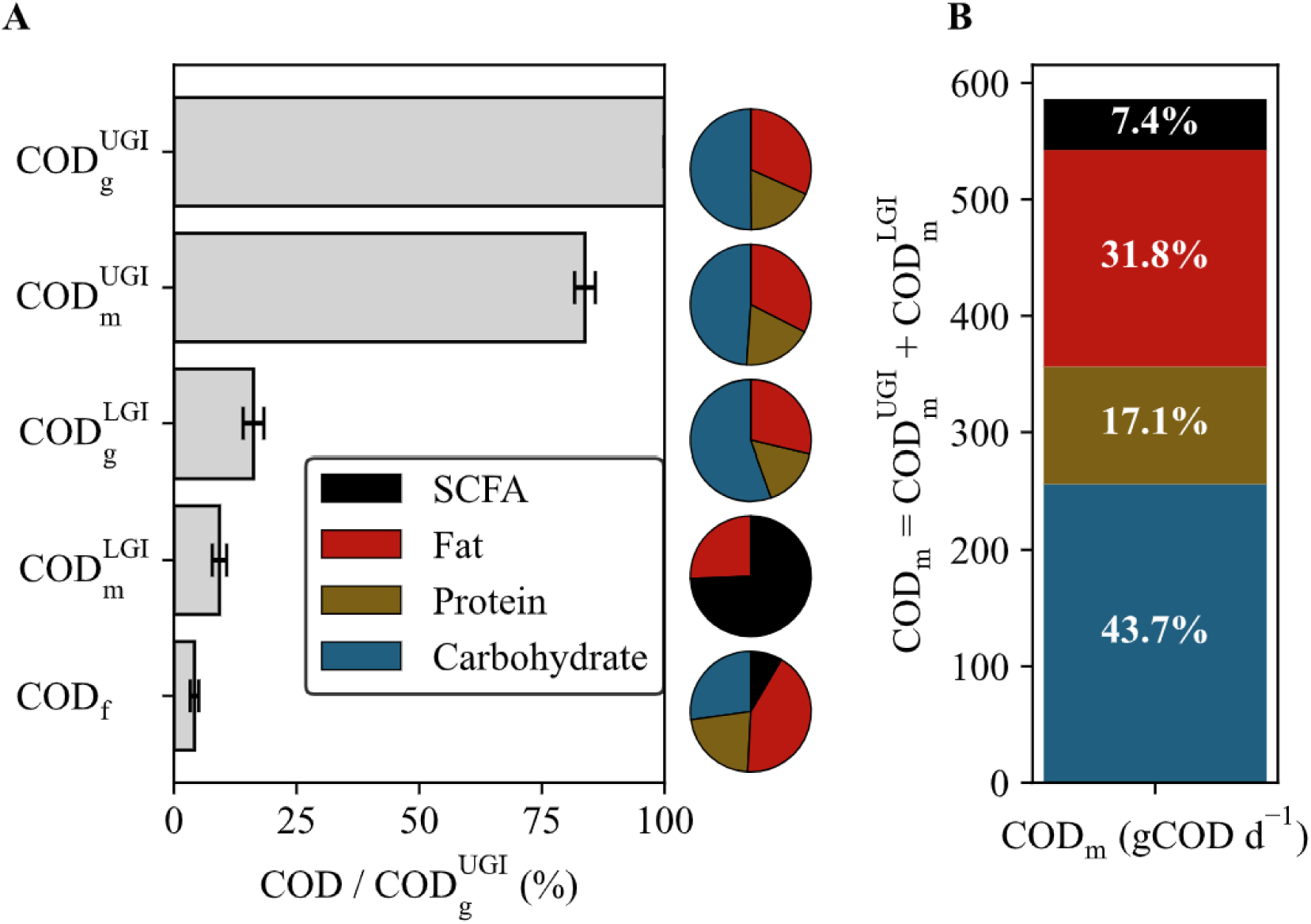
Mass balances for chemical oxygen demand (COD) in upper and lower gastrointestinal (UGI and LGI) tracts. **A)** The percent of the dietary intake (%) that each input and output stream of the UGI and LGI compartments within the Digestion partition of the DAMM model. Bars represent the average ± standard deviation for both diets. The top three bars - *COD_g_^UGI^*, *COD ^UGI^*, and *COD_g_^LGI^* - are the inputs/outputs for the UGI, while the bottom three bars - *COD_g_^LGI^, COD ^LGI^*, and *COD_f_* - are the inputs/outputs for the LGI. The fractions of SCFA, Fat, Protein, and Carbohydrate for each bar are shown on the pie charts directly to the right. **B)** A stacked bar chart of the average amounts of SCFA, Fat, Protein, and Carbohydrates within the DAMM predicted total metabolizable COD. Percentages of the total are listed on each bar segment. Subscripts for COD refer to the following: g – gross, m – metabolizable, and f – fecal.

Figure 2B combines the metabolizable COD (COD_m_) from the UGI and the LGI, and it segments the bars by proteins, carbohydrates, fats, and SCFAs. The model predicts an average COD_m_ of 93.0 ± 4.1%, within which approximately 85% came from the UGI and 15% from the LGI. SCFAs absorbed from the LGI contribute an average of 43.6 ± 12.0 gCOD d^-1^ to the metabolizable COD; this is 7.4% of the total COD_m_ and roughly equivalent to 140 kcal d^-1^. While protein, carbohydrate, and fat absorption in the UGI largely occurs with specific transporters after being broken down by enzymes (50), absorption in the LGI occurs after fermentation to products such as microbially produced SCFAs (51). By separating UGI and LGI absorptions, the DAMM model accounts for the distinct pathways for metabolizable COD; the prediction of microbial fermentation products in the LGI means that in the future, this information can be separately input into existing models that predict changes in body energy stores (52) and allow us to consider the microbial contribution explicitly.

### DAMM model captures interindividual variation of fecal SCFAs and biomass on diets

Inputs from a prior clinical trial (22) were used to generate DAMM model outputs. In that trial, two diets were administered in random order to each participant (crossover study design). Both diets had similar percentages of proteins, carbohydrates, and fats, but the MBD was designed to encourage fermentation in the LGI by having a higher amount of carbohydrates that are not digestible by the human host than the WD. Figure 3 depicts the DAMM model’s predicted metabolizable COD from total absorbed SCFAs, acetate, butyrate, and propionate. The predictions for each diet were significantly different, mirroring the directionality that was observed in the clinical trial (higher SCFAs in serum and feces on the MBD as compared to the WD). The butyrate values were the most similar, having a difference of only 3.3 gCOD d^-1^, which is roughly 10 kcals d^-1^. The average total predicted metabolizable COD from SCFAs was 51.9 ± 9.8 gCOD d^-1^ on the MBD and 33.8 ± 4.4 gCOD d^-1^ on the WD, which are 5% and 10% of the total metabolizable COD, respectively.

**Figure 3:**
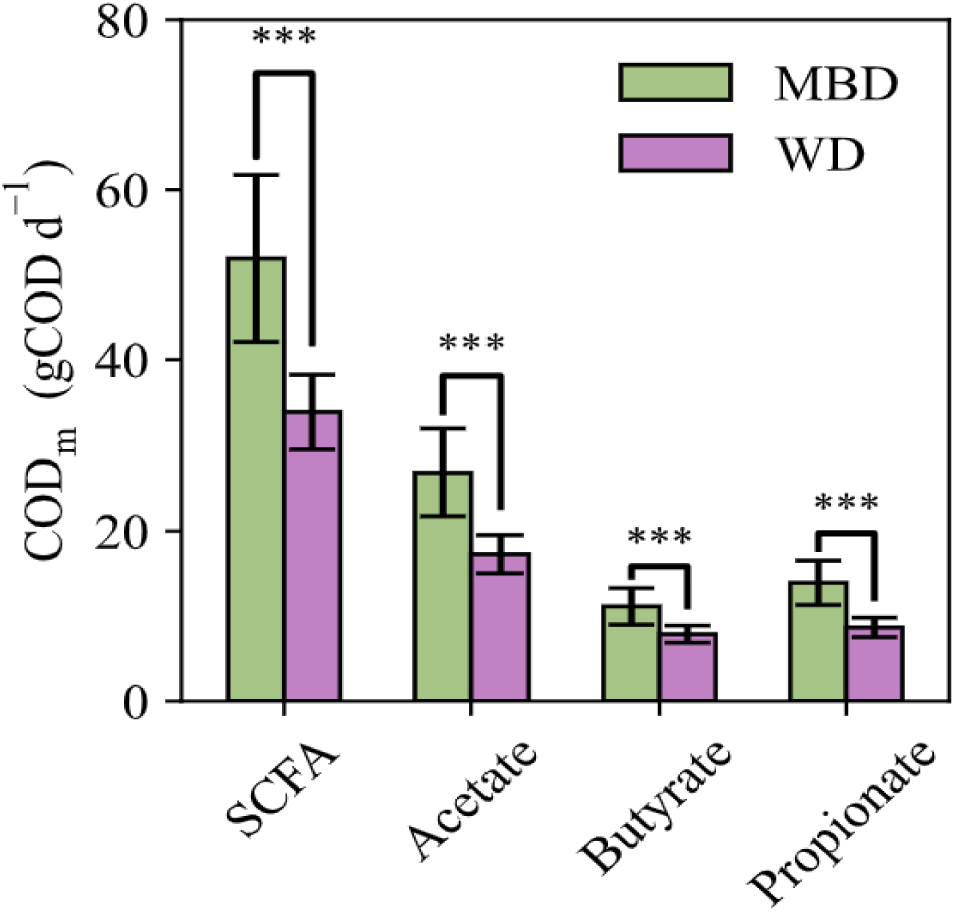
Predictions for short-chain fatty acid (SCFA) metabolizable chemical oxygen demand (COD_m_) by diet. Bars represent the mean ± standard deviation of all participants for each diet. Statistical significance was determined using a t-test: “***” indicates p < 0.001. Brackets above the bars highlight the comparisons between the two diets. Comparisons were performed for acetate, butyrate, propionate, and SCFA which is the sum of the three.

We cannot compare these predictions to measurements, because the clinical study could not directly measure SCFAs absorbed in the colon; however, the study measured the following SCFAs in the feces: acetate, propionate, and butyrate. The SCFA samples were from the 6-day composite fecal sample and the measurements were converted to the daily fecal COD (22). Figures 4A and **4B** compare the DAMM model’s daily fecal COD predictions for acetate, butyrate, and propionate for all participants versus the measurements. Apart from butyrate on the MBD, the predictions for SCFA fecal COD have similar scale and variation as the measurements. T-tests were run for each measurement and prediction pair, and all had p-values greater than 0.3, except for MBD butyrate where the p = 0.07 (Figure 4A).

**Figure 4:**
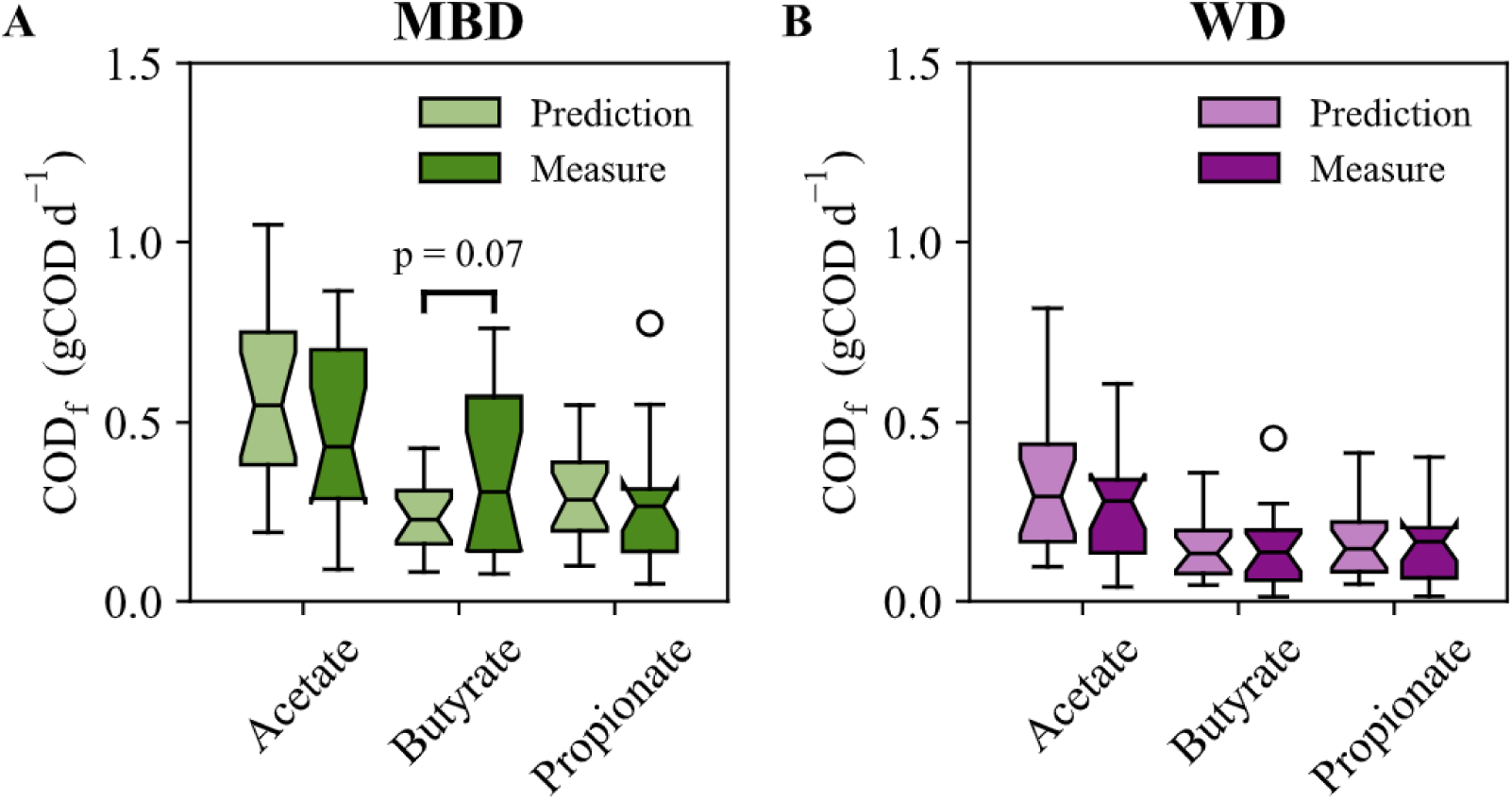
Daily fecal short-chain fatty acid (SCFA) chemical oxygen demand (COD) boxplots comparing predictions and measurements by diet. **A)** Predictions for acetate, butyrate, and propionate fecal COD directly compared to the measurements for the microbial enhancer diet (MBD). **B)** Predictions for acetate, butyrate, and propionate fecal COD directly compared to the measurements for the western diet (WD). Statistical significance was determined using a t-test. All p-values were greater than 0.3, except when labeled on the plot.

Standard error of estimates (σ_est_), relative to the 1:1 identity line, are listed in **Table 3** for acetate, butyrate, and propionate for the MBD and WD. The largest σ_est_, 0.27 gCOD d^-1^ for butyrate on the MBD, is equivalent to 0.9 kcal d^-1^; thus, the DAMM model predicted fecal SCFAs within one kilocalorie per day. The good agreement between the fecal SCFA predictions and measured values, in magnitude and trend, supports the validity of the SCFA predictions from the DAMM model. Explicitly including SCFA production and absorption rates was key to the model’s success. This is underscored by the fact that including CTT allowed the DAMM model to capture inter-individual variations in fecal-SCFA excretion rates; using a constant SCFA absorption ratio (e.g., 95%, as in the prior model (27)), would have yielded fecal SCFAs that spanned a range of less than 0.2 gCOD d^-1^ which we show in **Supplemental Material** Figure 2. While several factors can affect SCFA absorption (51,53,54), the accuracy of the DAMM predictions when using the CTT dependent absorption function show that CTT is an important factor.

In addition to the SCFAs, the model also tracked the microbial growth that resulted from the defined reaction categories: carbohydrate fermentation, protein fermentation, and methanogenesis. Previous analyses on 16S rRNA gene qPCR measurements demonstrated a statistically significant increase in fecal biomass on the MBD as compared to the WD (31). As shown in Figure 5A, the model accurately captured this observation. DAMM predicted an increase of 50 mgCOD for every gram of substrate COD that enters the colon on the MBD, compared with the WD.

**Figure 5:**
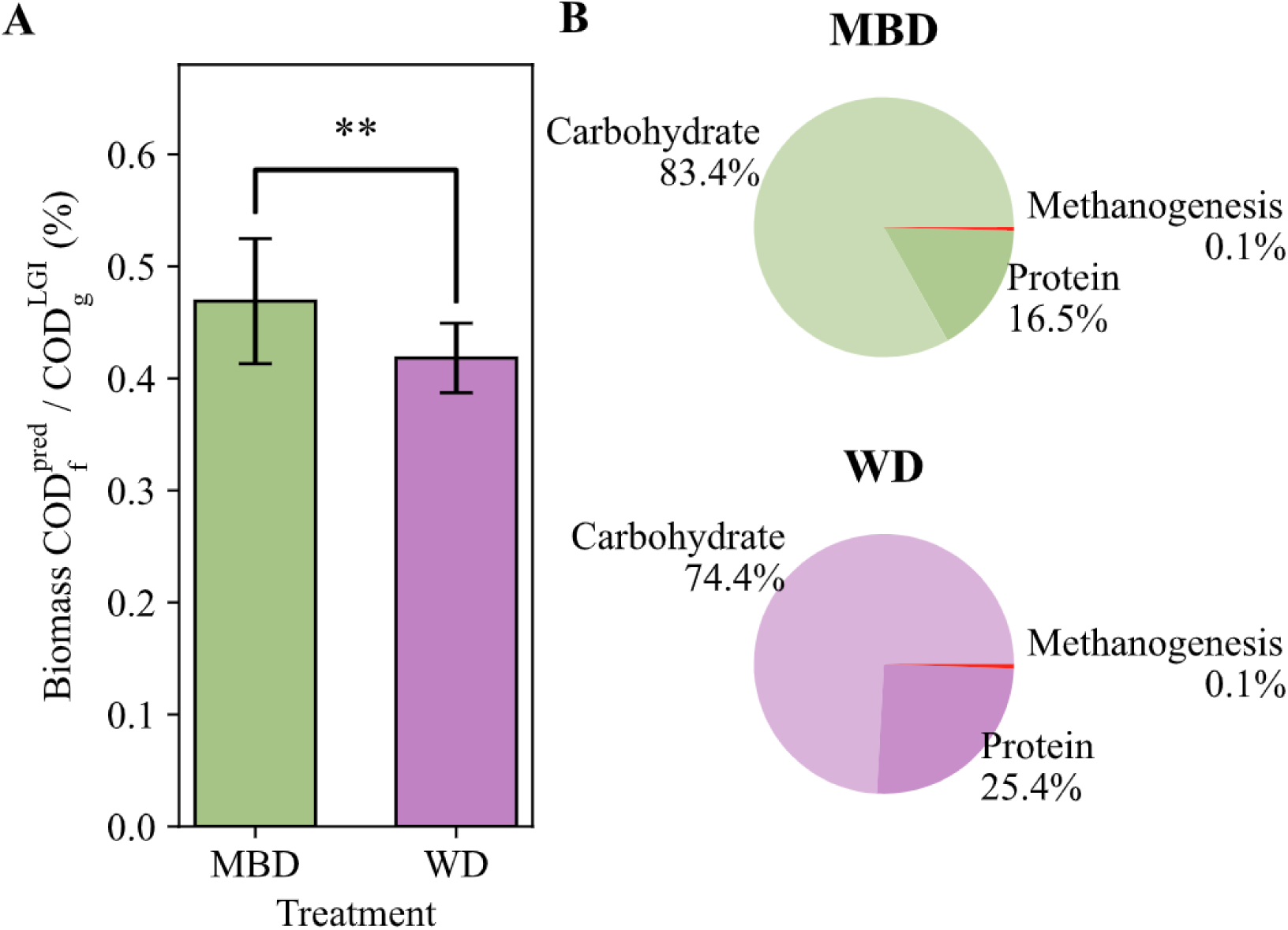
Fecal biomass chemical oxygen demand (COD) comparison between dietary treatments. **A)** Direct comparison of the predicted fecal biomass COD (Biomass COD_f_ ^pred^) relative to the substrates COD coming into the colon (COD_g_^LGI^) on each diet. Bars represent the mean ± standard deviation of all participants for each diet. Statistical significance was determined using a t-test where p = 0.002. The “**” on the chart indicates p < 0.01. **B)** Pie charts display the fraction of predicted fecal biomass that is attributed to each of the three reaction categories: carbohydrate fermentation, protein fermentation, and methanogenesis for each diet. MBD is the microbiome enhancer diet, and WD is the western diet.

Figure 5B illustrates the average proportion of biomass attributed to each of the three reaction categories. For both diets, most of the biomass was from carbohydrate and protein fermentation, while biomass from methanogenesis was very small; methanogens are discussed more in the next section. Due to the higher amount of fiber and resistant starch on the MBD, which were not absorbed in the upper gastrointestinal tract, the percentage of fecal biomass from carbohydrate fermentation was 10% higher for the MBD treatment compared to the WD. The greater availability of fiber-derived carbohydrates in the lower gastrointestinal tract resulted in increased carbohydrate fermentation by the microbial community.

### Methane predictions support hypothesis that methanogens are present in biofilms in colon

The prior clinical trial measured fecal-methanogen abundance using qPCR targeting the *mcrA* gene (55), and we used these data to provide an initial estimate of methanogen biomass (X_M_^0^) for each participant (**Supplemental Material Table 1**). This X_M_^0^ was an input into the DAMM model to predict daily methane COD production (COD_CH4_).

In Figure 6A, we compare the methane predictions with the measurements from the clinical trial for participants who produced significant methane (i.e., methane measurements exceeded 100 mL d^-1^) (31,46). The predictions differ by at least an order of magnitude from the measurements for methane-producing participants. We calculated the standard error of the estimate (σ_est_) relative to the identity line as 0.05 gCOD d^-1^, with a coefficient of determination (R^2^) of −278%. The very large systematic deviation in the model’s predictions indicates that our initial X_M_ ^0^ estimates were too small. This conclusion is reinforced by Figure 5B, in which the fraction of methanogenesis was negligible compared to those for protein and carbohydrate fermentation.

**Figure 6:**
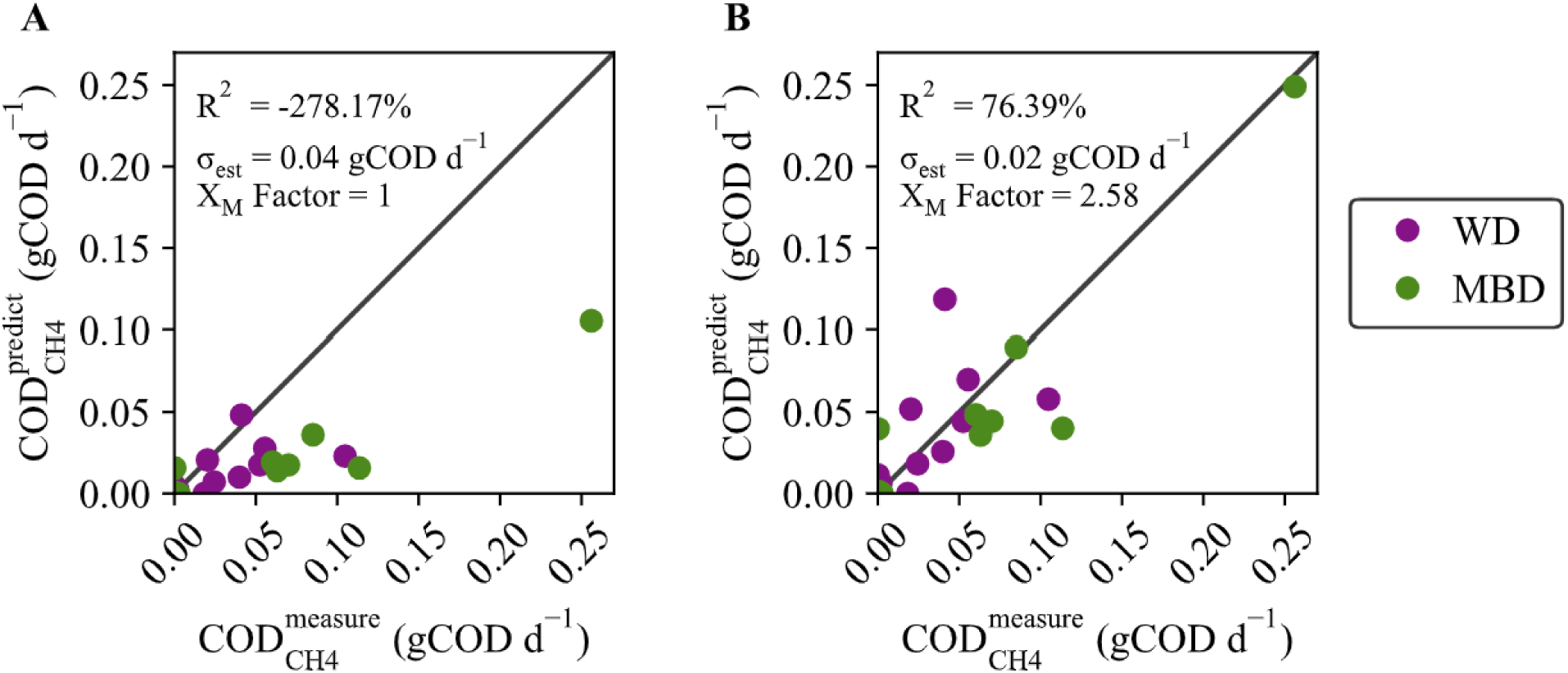
Predicted daily methane production in chemical oxygen demand (COD) compared to measurements for participants who generated measured methane. **A)** Comparing daily methane production (COD_CH4_^predict^) predictions to the (COD_CH4_^measure^) measurement when the initial methanogen biomass (X_M_^0^) is calculated directly from the *mcrA* qPCR copy numbers (X_M_ Factor = 1). R^2^ for the identity line is −278.17% and σ_est_ = 0.04 gCOD d^-1^. **B)** Comparing COD^predict^ to the COD measure when X is proportional to the *mcrA* qPCR copy numbers times the factor 2.58 (X_M_ Factor = 2.58). R^2^ for the identity line is 76.39% and σ_est_ = 0.02 gCOD d^-1^. Data excludes one outlier where methanogens were measured in the feces, but the participant had minimal methane production. Symbol colors are by diet, where WD is the western diet and MBD is the microbiome enhancer diet.

The likely explanation for the systematic low values in Figure 6A is that most of the methanogens present in the colon were not in the feces, but were accumulated in a biofilm on the intestine’s epithelium. We evaluated this hypothesis by assuming that the true mass of methanogens was proportional to the methanogens measured in the feces, but larger by a solids-concentration factor (*X*_M_ Factor) that reflected that the methanogens had a larger SRT than the feces’ CTT.

We adapted Equation 2 to include the *XM* Factor to account for the accumulation of methanogens as a biofilm in the epithelium.

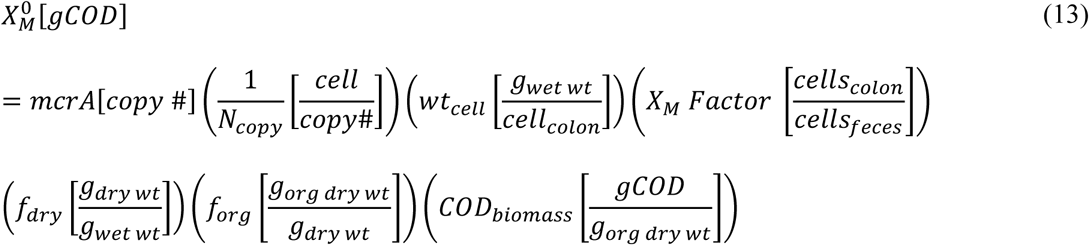

We used a grid-search algorithm that minimized the σ_est_ and calculated the optimum *X_M_* Factor, which was 2.58 cell_colon_ cell_feces_^-1^. Figure 6B shows that using a constant *X_M_* Factor put all predicted values in the correct order of magnitude and eliminated systematic bias.

Allowing the methanogens to have a longer SRT within the colon dramatically improved results, which points to methanogens being retained in a biofilm in the mucus layer of the colon. Biofilms provide a stable environment in which methanogens, being slow growers, can thrive despite the relatively short CTT (1 – 2 days) (11). *Methanobrevibacter smithii*, the most common methanogen archaea in the human feces (31), has been shown capable of forming biofilms on different substrates (56,57), and it can also be retained in a biofilm formed by other microorganisms or by epithelium mucus (58). Having the methanogens in an epithelium biofilm would accentuate their interactions with the human host, including processing and absorption of SCFAs. Some evidence indicates that microorganisms in epithelial biofilms influence bowel inflammation and dysbiosis (59,60), while methanogens, specifically, *M. smithii*, have been linked to obesity (61) and anorexia (62). Further research is necessary, but the DAMM model creates a starting point to expand our understanding of how methanogens might contribute to health or disease.

### DAMM model improves predictions for metabolizable chemical oxygen demand

The DAMM model estimates the metabolizable COD_m_ (which can be directly converted to metabolizable energy (12)) from the upper and lower gastrointestinal tracts. We compared the performance of the DAMM to the standard Atwater model (15) for predicting the metabolizable COD of all participants in the clinical study (22). Figure 7 illustrates the predictive performance of the DAMM model (Figures 7A and **7C**) and the Atwater constants (Figures 7B and **7D**) compared to the actual COD_m_ (calculated as the difference between dietary intake and measured fecal COD). In Figure 7A, the DAMM model more accurately predicted the COD_m_, as shown by the good fit to the identity line and the statistical metrics: R^2^ = 95.6% and σ_est_ = 20.6 gCOD d^-1^. The DAMM results are an improvement over the Atwater predictions (Figure 7C), which consistently underestimated COD_m_ and had larger errors: R^2^ = 88.5% and σ_est_ = 32.6 gCOD d^-1^.

**Figure 7:**
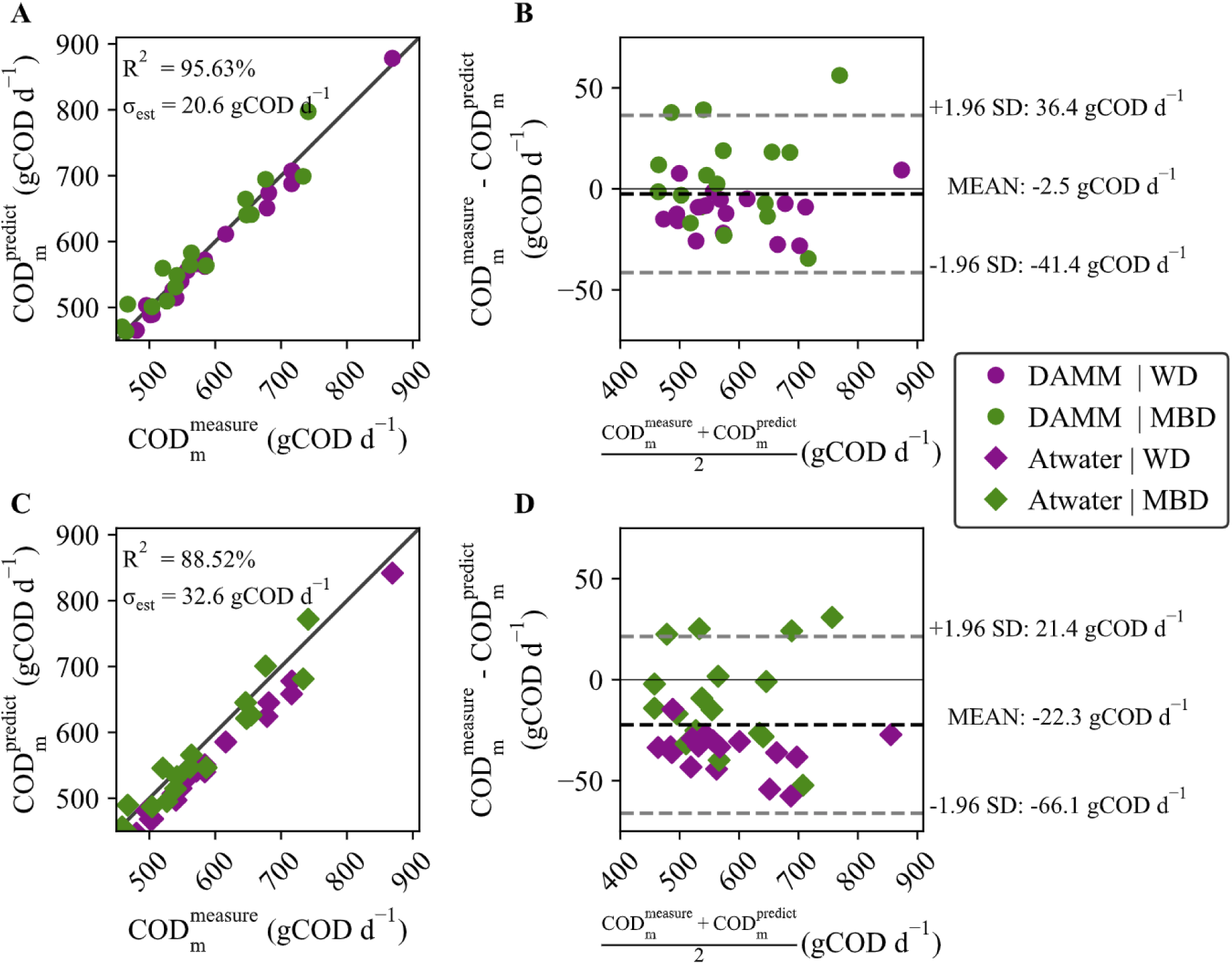
Accuracy and bias evaluation of DAMM and Atwater metabolizable chemical oxygen demand (CODm) predictions. **A)** Scatterplot of DAMM prediction versus measurement. The coefficient of determination (R^2^) and standard error of estimate (σ_est_) for the identity line are 95.63% and 20.6 gCOD d^-1^. **B)** Bland-Altman plot showing the 95% confidence intervals at 36.4 gCOD d^-1^ to −41.4 gCOD d^-1^ with a mean of 2.5 gCODd^-1^ for the measurements and DAMM predictions. **C)** Scatterplot of Atwater prediction versus measurement. R^2^ and σ_est_ for the identity line are 88.52% and 32.6 gCOD d^-1^. **D)** Bland-Altman plot showing the 95% confidence intervals at 21.4 gCOD d^-1^ to −66.1 gCOD d^-1^ with a mean of −22.3 gCOD d^-1^ for the measurements and Atwater-constant predictions.

**Table 4** quantifies similar differences when each diet is considered separately.

**Table 4:** Metabolizable chemical oxygen demand (CODm) statistical metrics. Includes coefficient of determination (R^2^), standard error of estimate (σest), one sample t-test against the difference of the methods (t-test), and Kendall’s coefficient of concordance (τ) to compare predictions to measurements for DAMM and Atwater models.

| Model | Treatment | $R^2$ (%) | $\sigma_{\text{est}}$ (gCOD d<sup>-1</sup>) | t-test (gCOD d<sup>-1</sup>) | $\tau$ |
| --- | --- | --- | --- | --- | --- |
| DAMM | WD | 92.87% | 16.23 | 0.98<br>(p = 0.34) | -0.08<br>(p = 0.66) |
|  | MBD | 97.78% | 25.38 | -0.24<br>(p = 0.81) | 0.03<br>(p = 0.90) |
|  | COMBINED | 95.63% | 20.62 | 0.12<br>(p = 0.90) | 0.05<br>(p = 0.65) |
| Atwater | WD | 85.75% | 39.14 | 3.37<br>(p = 0.003) | 0.19<br>(p = 0.31) |
|  | MBD | 91.84% | 27.00 | 0.37<br>(p = 0.71) | 0.03<br>(p = 0.90) |
|  | COMBINED | 88.52% | 32.55 | 0.97<br>(p = 0.34) | 0.13<br>(p = 0.27) |

The Bland-Altman plots for the DAMM (Figure 7B) and Atwater constants (Figure 7D) further highlight the improvement in accuracy with the DAMM: The mean differences between predictions and measurements were only −2.5 gCOD d^-1^ for the DAMM, compared to −22.3 gCOD d^-1^ for the Atwater constants, although the limits of agreement are roughly the same at approximately ± 40 gCOD d^-1^. The Bland-Altman plots also show that, while the MBD treatment was evenly distributed, the WD clustered below the mean difference for both models.

We conducted a one-sample t-test to assess systematic bias and calculated Kendall’s coefficient of concordance (τ) to examine proportional bias for each diet individually, as well as for the combined dataset that includes both treatments. Only the Atwater WD predictions had statistically significant bias: t-test = 3.37 gCOD d^-1^ and p = 0.002 (**Table 4**). According to the τ calculations for the mean difference and average of the measurements and predictions (**Table 4**), neither of the models had a proportional bias.

The improvement in COD_m_ predictions by the DAMM model, compared to the commonly used Atwater factors (15), demonstrates that the DAMM model is a promising alternative to the current Atwater approach. One of the primary advantages of using the DAMM model is that it provides added insight by distinguishing between the upper and lower gastrointestinal tracts and explicitly incorporating microbial metabolism. This contrasts to the traditional Atwater approach, which treats the entire digestive system as a “black box.” We show that the Atwater factors had a systematic bias of consistently underpredicting metabolizable COD_m_, particularly for the WD. The source of the bias could be a *de facto* underestimation of SCFA absorption on diets that contain fewer complex carbohydrates. By explicitly including SCFA production and absorption, the DAMM model more accurately captured the microbial contribution.

## Conclusion

Microorganisms play a fundamental role in human digestion and are essential for colonic health (63), but standard methods of calculating metabolizable energy (15) do not provide any insight into the composition and metabolic functioning of the colon’s microbial community. The DAMM model overcomes this limitation, because it predicts the structure and function of the colon’s microbial community for a specific diet. The DAMM model provides a mechanistic framework that divides the digestive tract into two major physiological sections and explicitly includes microbial metabolism. DAMM model predictions allow us to know if a diet is starving or adequately feeding the colon’s microbial community.

By representing the structure and metabolic function of the colon’s microbial community, the DAMM model 1) introduced an opportunity to develop human metabolic models that incorporate absorption of microbially produces SCFAs in the future; 2) captured observed differences between the two diets in terms microbially derived SCFAs, fecal COD, and in metabolizable COD; 3) pointed to methanogens accumulating in epithelial biofilms which tells us more about possible community structure and improved methane predictions; and 4) reduced systematic bias inherent in the Atwater model, which does not explicitly include the microbiome.

## Acronym Table

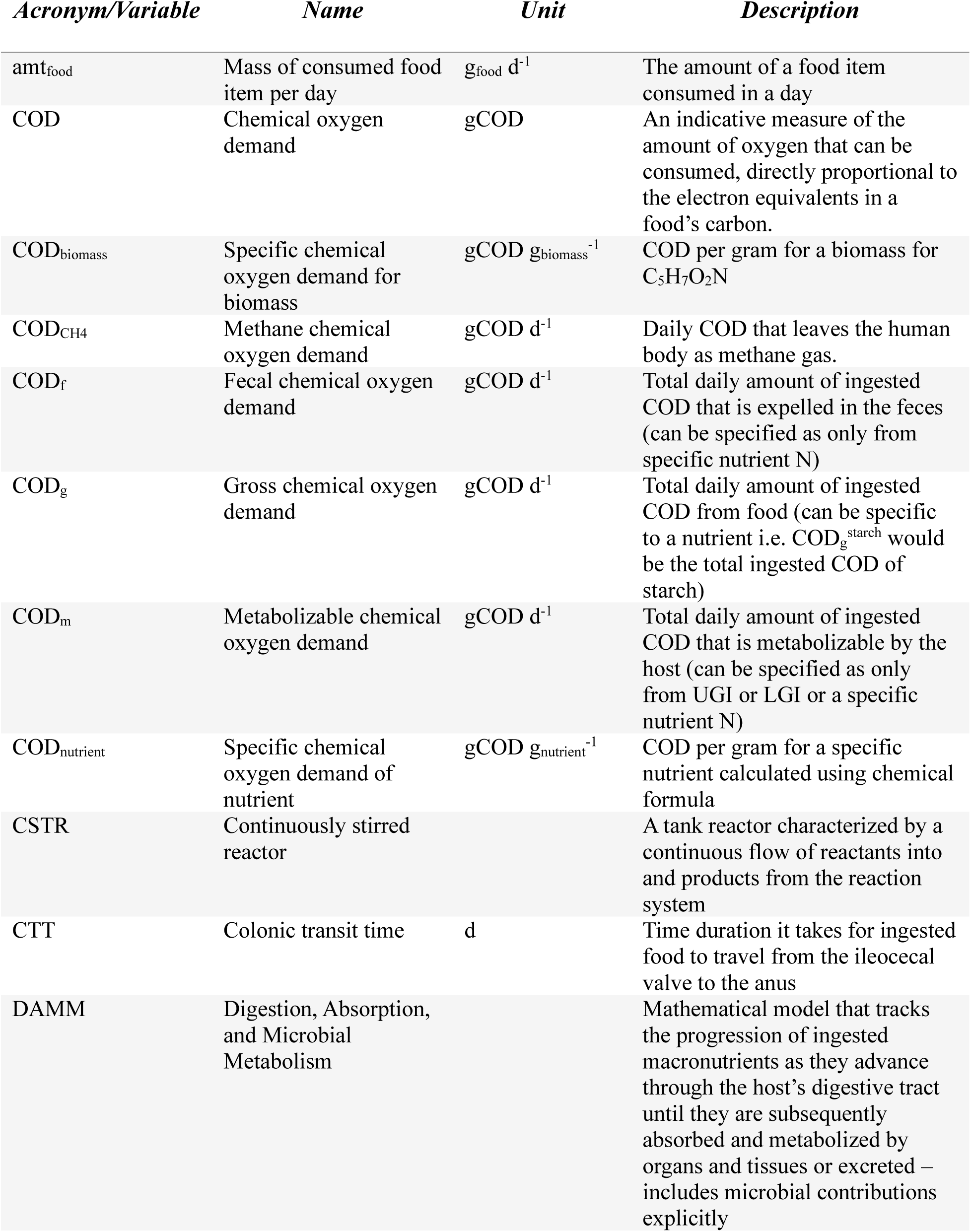
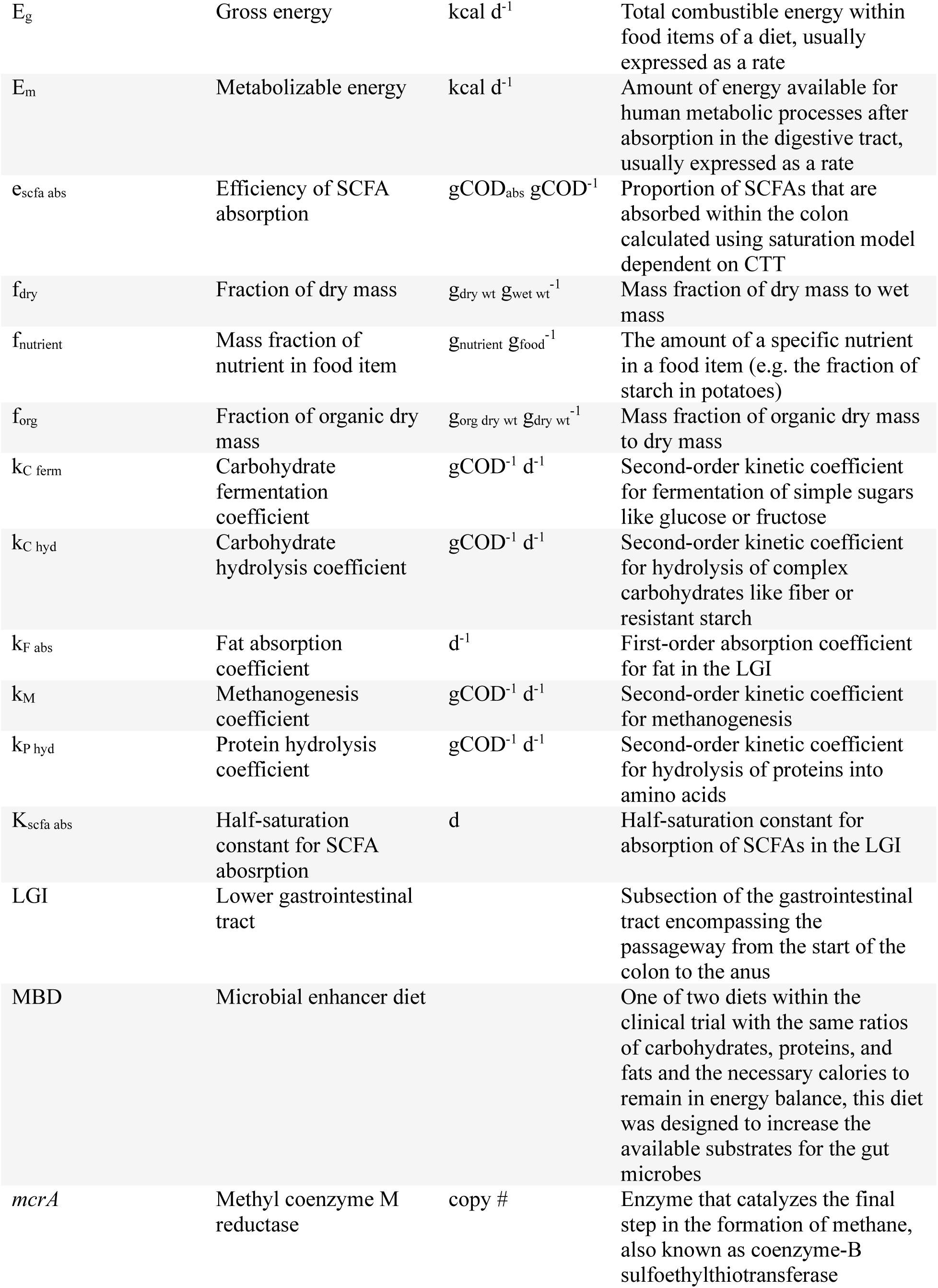
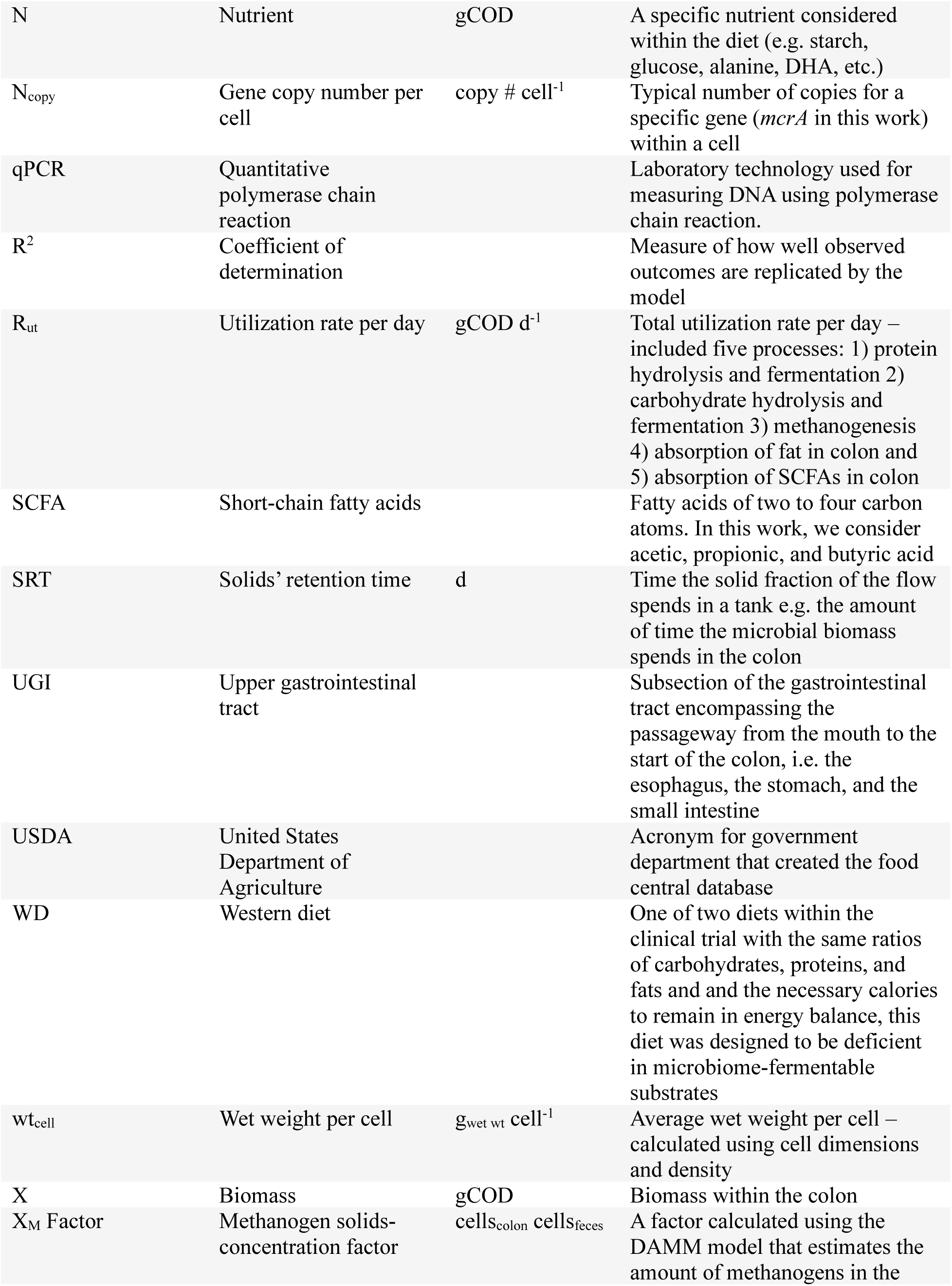
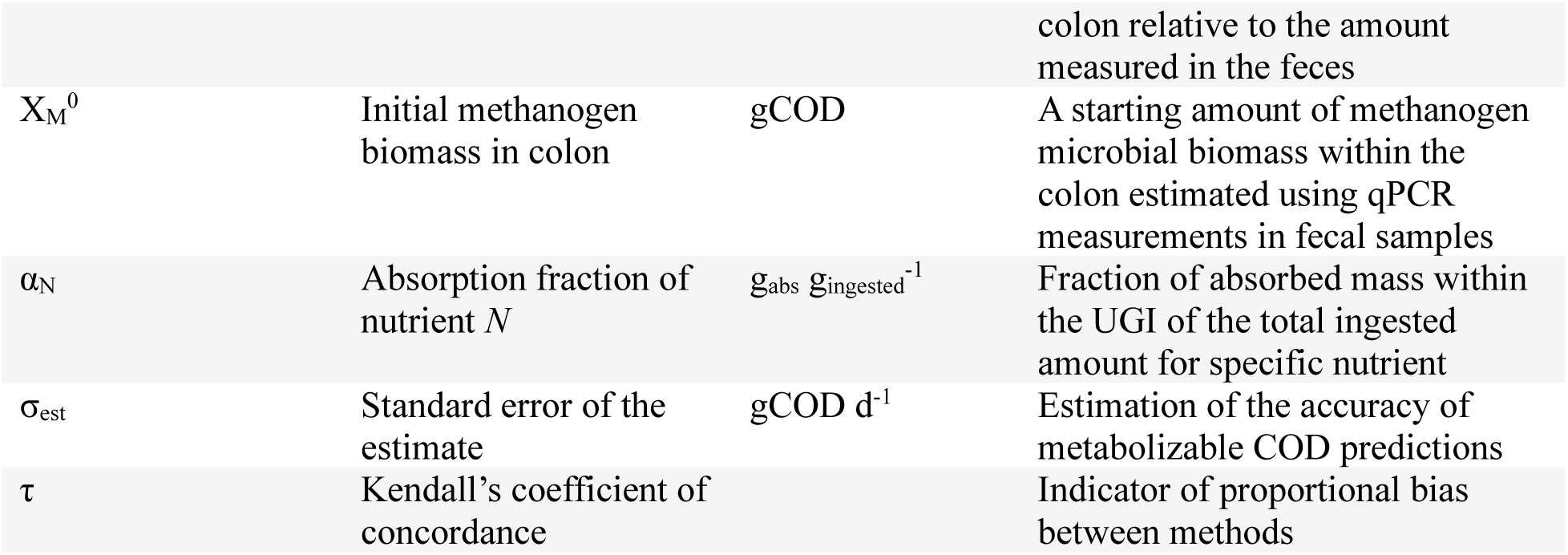

## Data Availability

All data produced in the present study are available upon reasonable request to the authors.

