## Supplemental Material for "Modeling the microbial contribution to human Energy Balance using the Digestion, Absorption, and Microbial Metabolism (DAMM) model"

### Supplemental Material Tables

**Supplemental Material Table 1:** DAMM inputs for measured colonic transit time (CTT) (1) and calculated initial methanogen biomass concentration from *mcrA* qPCR measurements (2). The starred value is a removed outlier where minimal methane was measured despite the presence of methanogens within the feces.

| ID (Treatment) | $X_M^0$ [$10^3$ gCOD] | CTT [d] |
|---|---|---|
| P01 (MBD) | 1.9 | 0.9 |
| P01 (WD) | 0.8 | 0.9 |
| P02 (MBD) | 0 | 3.0 |
| P02 (WD) | 0 | 1.6 |
| P03 (MBD) | 2.7 | 0.9 |
| P03 (WD) | 1.3 | 2.4 |
| P04 (MBD) | 0 | 1.5 |
| P04 (WD) | 0.1 | 1.0 |
| P05 (MBD) | 2.2 | 1.0 |
| P05 (WD) | 1.3 | 3.9 |
| P06 (MBD) | 0.1 | 2.7 |
| P06 (WD) | 0 | 3.8 |
| P07 (MBD) | 0 | 0.9 |
| P07 (WD) | 0 | 1.4 |
| P08 (MBD) | 0 | 0.7 |
| P08 (WD) | 0 | 1.2 |
| P09 (MBD) | 8.2 | 1.4 |
| P09 (WD) | 3.4 | 2.5 |
| P10 (MBD) | 0 | 1.0 |
| P10 (WD) | 0 | 1.2 |
| P11 (MBD) | 2.0 | 1.7 |
| P11 (WD) | 1.8 | 2.7 |
| P12 (MBD) | 3.6 | 1.9 |
| P12 (WD) | 0.6 | 0.7 |
| P13 (MBD) | 0.8* | 0.5 |
| P13 (WD) | 1.7 | 0.5 |
| P14 (MBD) | 0 | 0.83 |
| P14 (WD) | 0 | 0.9 |
| P15 (MBD) | 0 | 0.6 |
| P15 (WD) | 0 | 0.7 |

| | | |
|---|---|---|
| P16 (MBD) | 0 | 0.1 |
| P16 (WD) | 0 | 1.7 |
| P17 (MBD) | 0 | 1.1 |
| P17 (WD) | 0 | 0.6 |

**Supplemental Material Table 2:** DAMM digestivity constants for absorption within the UGI.

| Macronutrient | Nutrient | α | Source |
|---|---|---|---|
| *carbohydrates* | Sucrose | 0.975 | (3) |
| | Glucose | 0.975 | |
| | Fructose | 0.975 | |
| | Lactose | 0.975 | |
| | Maltose | 0.975 | |
| | Galactose | 0.975 | |
| | Starch | 0.925 | |
| | Cellulose | 0.08 | (4) |
| *fats* | Butyric acid | 1.000 | (5) |
| | Caproic acid | 1.000 | |
| | Caprylic acid | 0.000 | |
| | Capric acid | 0.974 | |
| | Lauric acid | 0.873 | |
| | Myristic acid | 0.778 | |
| | Palmitic acid | 0.208 | |
| | Stearic acid | 0.000 | |
| | Arachidic acid | 0.000 | |
| | Behenic acid | 0.767 | |
| | Palmitoleic acid | 0.939 | |
| | Oleic acid | 0.000 | |
| | Linoleic acid | 0.812 | |
| | α-Linolenic acid | 0.985 | |
| | Arachidonic acid | 0.799 | |
| | EPA | 0.359 | |
| | DPA | 0.000 | |
| | DHA | 0.000 | |
| *proteins* | Alanine | 0.881 | (4) |
| | Arginine | 0.902 | |
| | Asparagine | 0.873 | |
| | Cystine | 0.855 | |
| | Glutamine | 0.936 | |
| | Glycine | 0.715 | |
| | Histidine | 0.902 | |
| | Isoleucine | 0.909 | |
| | Leucine | 0.919 | |
| | Lysine | 0.936 | |
| | Methionine | 0.931 | |
| | Phenylalanine | 0.896 | |
| | Proline | 0.899 | |

| | |
|---|---|
| Serine | 0.865 |
| Threonine | 0.847 |
| Tryptophan | 0.767 |
| Tyrosine | 0.889 |
| Valine | 0.897 |

**Supplemental Material Table 3:** Stoichiometry matrix for the hydrolysis and fermentation of the carbohydrates and proteins within the LGI tract compartment. The matrix units are in gCOD unless specified as grams of nitrogen (gN) instead. Valerate is excluded v from this table, but it is present in the stoichiometry matrix.

| *Electron donors:* | *Donor* | *Microbial biomass* | *Hydrogen sulfide* | *Ammonia (gN)* | *Acetate* | *Propionate* | *Butyrate* | *Methane* | *Hydrogen* |
|---|---|---|---|---|---|---|---|---|---|
| *Unspecified Carb* | -1.00 | 0.20 | 0.00 | -0.02 | 0.42 | 0.23 | 0.16 | 0.00 | 0.00 |
| *Sucrose* | -1.00 | 0.20 | 0.00 | -0.02 | 0.42 | 0.23 | 0.16 | 0.00 | 0.00 |
| *Glucose (dextrose)* | -1.00 | 0.20 | 0.00 | -0.02 | 0.42 | 0.23 | 0.16 | 0.00 | 0.00 |
| *Fructose* | -1.00 | 0.20 | 0.00 | -0.02 | 0.42 | 0.23 | 0.16 | 0.00 | 0.00 |
| *Lactose* | -1.00 | 0.20 | 0.00 | -0.02 | 0.42 | 0.23 | 0.16 | 0.00 | 0.00 |
| *Maltose* | -1.00 | 0.20 | 0.00 | -0.02 | 0.42 | 0.23 | 0.16 | 0.00 | 0.00 |
| *Galactose* | -1.00 | 0.20 | 0.00 | -0.02 | 0.42 | 0.23 | 0.16 | 0.00 | 0.00 |
| *Starch* | -1.00 | 0.20 | 0.00 | -0.02 | 0.42 | 0.23 | 0.16 | 0.00 | 0.00 |
| *Fiber, total dietary* | -1.00 | 0.20 | 0.00 | -0.02 | 0.42 | 0.23 | 0.16 | 0.00 | 0.00 |
| *Unspecified Protein* | -1.00 | 0.20 | 0.00 | 0.11 | 0.35 | 0.09 | 0.35 | 0.00 | 0.00 |
| *Alanine* | -1.00 | 0.20 | 0.00 | 0.13 | 0.00 | 0.00 | 0.00 | 0.00 | 0.80 |
| *Arginine* | -1.00 | 0.22 | 0.00 | 0.30 | 0.15 | 0.25 | 0.00 | 0.00 | -0.09 |
| *Aspartic acid* | -1.00 | 0.20 | 0.00 | 0.20 | 0.53 | 0.00 | 0.00 | 0.00 | 0.27 |
| *Cystine* | -1.00 | 0.17 | 0.17 | 0.13 | 0.59 | 0.00 | 0.00 | 0.00 | 0.07 |
| *Glutamic acid* | -1.00 | 0.20 | 0.00 | 0.18 | 0.36 | 0.00 | 0.44 | 0.00 | 0.00 |
| *Glycine* | -1.00 | 0.27 | 0.00 | 0.27 | 1.07 | 0.00 | 0.00 | 0.00 | -0.33 |
| *Histidine* | -1.00 | 0.20 | 0.00 | 0.25 | 0.36 | 0.00 | 0.44 | 0.00 | 0.00 |

| | | | | | | | | | |
|---|---|---|---|---|---|---|---|---|---|
| *Isoleucine* | -1.00 | 0.20 | 0.00 | 0.04 | 0.00 | 0.00 | 0.00 | 0.00 | 0.11 |
| *Leucine* | -1.00 | 0.20 | 0.00 | 0.04 | 0.00 | 0.00 | 0.00 | 0.00 | 0.11 |
| *Lysine* | -1.00 | 0.20 | 0.00 | 0.11 | 0.23 | 0.00 | 0.57 | 0.00 | 0.00 |
| *Methionine* | -1.00 | 0.18 | 0.08 | 0.06 | 0.00 | 0.64 | 0.00 | 0.00 | 0.09 |
| *Phenylalanine* | -1.00 | 0.20 | 0.00 | 0.03 | 0.00 | 0.00 | 0.00 | 0.00 | 0.80 |
| *Proline* | -1.00 | 0.22 | 0.00 | 0.06 | 0.15 | 0.25 | 0.00 | 0.00 | -0.09 |
| *Serine* | -1.00 | 0.20 | 0.00 | 0.16 | 0.64 | 0.00 | 0.00 | 0.00 | 0.16 |
| *Threonine* | -1.00 | 0.22 | 0.00 | 0.09 | 0.40 | 0.00 | 0.50 | 0.00 | -0.13 |
| *Tryptophan* | -1.00 | 0.20 | 0.00 | 0.06 | 0.00 | 0.00 | 0.00 | 0.00 | 0.80 |
| *Tyrosine* | -1.00 | 0.20 | 0.00 | 0.03 | 0.00 | 0.70 | 0.00 | 0.00 | 0.10 |
| *Valine* | -1.00 | 0.20 | 0.00 | 0.06 | 0.00 | 0.00 | 0.67 | 0.00 | 0.13 |
| *Hydrogen* | | 1.02 | 0.00 | -0.01 | -1.02 | 0.00 | 0.00 | 1.00 | -1.00 |
| *Influent* | $Y_{inf}/\Theta_x$ | 0.00 | 0.00 | 0.00 | 0.00 | 0.00 | 0.00 | 0.00 | 0.00 |
| *Effluent* | $-Y_{eff}/\Theta_x$ | $-Y_{eff}/\Theta_x$ | $-Y_{eff}/\Theta_x$ | $-Y_{eff}/\Theta_x$ | $-Y_{eff}/\Theta_x$ | $-Y_{eff}/\Theta_x$ | $-Y_{eff}/\Theta_x$ | $-Y_{eff}/\Theta_x$ | $-Y_{eff}/\Theta_x$ |

**Supplemental Material Figures**

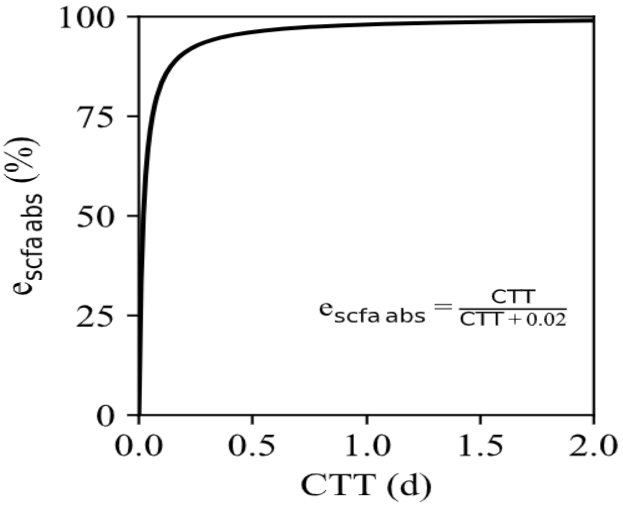


**Supplemental Material Figure 2: SCFA Absorption Saturation Model**

The saturation model (Eq. 12) for the absorption of SCFAs within the colon calculates the percentage of absorbed SCFAs for CTT up to 2 days.

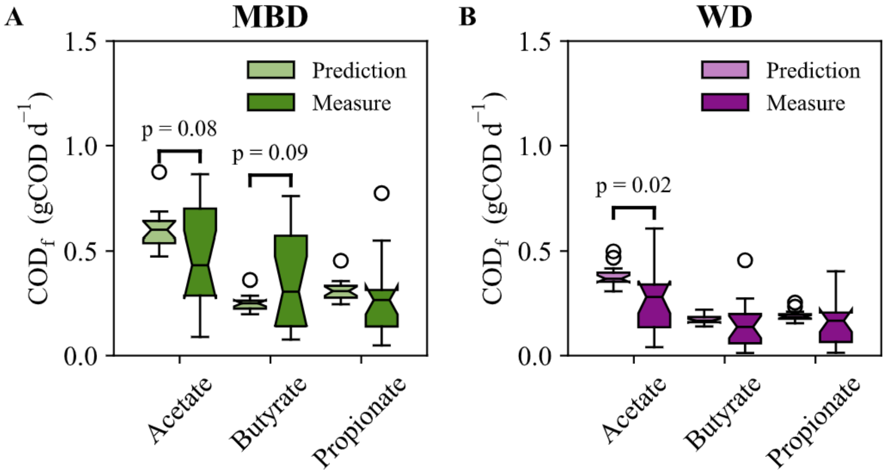


**Supplemental Material Figure 1: Predicted Fecal SCFAs with fixed 95% Absorption instead of Saturation Model**

A) Predictions for acetate, butyrate, and propionate fecal COD directly compared to the measurements for the microbial enhancer diet (MBD). B) Predictions for acetate, butyrate, and propionate fecal COD directly compared to the measurements for the western diet (WD). Statistical significance was determined using a t-test. All p-values > 0.3, except when labeled on the plot

### Supplemental Material Methods

### Methanogen Yield Calculations

We used the same half-reaction approach from Chapter 5 of Rittmann and McCarty (6) to develop the stoichiometry for all the other reactions as we did for methanogenesis; however, since we assumed the methanogens used acetate ($CH_3COOH$) as a carbon source for the cell synthesis, while hydrogen ($H_2$) was the electron donor for the energy production (7), the calculations were slightly more complicated and are summarized here.

The method balances half reactions for the electron donor for energy production ($H_2$), the carbon source for cell synthesis ($CH_3COOH$), the formation of microbial cells (methanogens – $C_5H_7O_2N$), and the electron acceptor ($CO_2$ reduced to $CH_4$) by systematically weighing the Gibb's free energy associated with the half reactions in Tables 5.3 to 5.5 in Rittmann and McCarty (6).

**$R_c^{synthesis}$:** $$\frac{1}{4}CO_2 + e^- + H^+ \rightarrow \frac{1}{4}H_2O + \frac{1}{8}CH_3COOH \qquad \Delta G_{cs}^{0}{}' = 27.4\frac{kJ}{e^-} \tag{3}$$

**$R_d^{energy}$:** $$e^- + H^+ \rightarrow \frac{1}{2}H_2 \qquad \Delta G_d^{0\prime} = 39.9\frac{kJ}{e^-} \tag{4}$$

**$R_{cells}$:** $$\frac{1}{20}NH_4^+ + \frac{1}{20}HCO_3^- + \frac{1}{5}CO_2 + e^- + H^+ \rightarrow \frac{9}{20}H_2O + \frac{1}{20}C_5H_7O_2N \qquad * \tag{5}$$

**$R_a$:** $$\frac{1}{8}CO_2 + e^- + H^+ \rightarrow \frac{1}{4}H_2O + \frac{1}{8}CH_4 \qquad \Delta G_a^{0\prime} = -23.5\frac{kJ}{e^-} \tag{6}$$

First, we calculated the energy required to convert the carbon source to activated acetate ($\Delta G^{0\prime} = 30.9$ kJ/$e^-$), the common organic intermediate that cells use in synthesis.

$$\Delta G_p = 30.9\left[\frac{kJ}{e^-}\right] - \Delta G_{cs}^{0\prime} = 30.9 - 27.4 = 3.5\frac{kJ}{e^-} \tag{1}$$

In cells, activated acetate is converted to cellular carbon, which has an estimated energy of $\Delta G_{pc} = 18.8\frac{kJ}{e^-}$ (6). We assumed the energy-transfer efficiency was $\varepsilon = 0.6$ We set $n = 1$ because additional energy is required to convert acetate to activated acetate. Cell synthesis energy requirement was then calculated using Supplemental Material Equation 2.

$$\Delta G_s = \frac{\Delta G_p}{\varepsilon^n} + \frac{\Delta G_{pc}}{\varepsilon} = \frac{3.5}{0.6^1} + \frac{18.8}{0.6} = 37.2\frac{kJ}{e^-} \quad (2)$$

Then, using the listed Gibb's free energy for the electron acceptor reaction, methane, and the electron donor for the energy production, hydrogen, we calculated the Gibb's energy of the reaction.

$$\Delta G_r = \Delta G_a^{0'} - \Delta G_d^{0'} = -23.5 - 39.9 = -63.4\frac{kJ}{e^-} \quad (7)$$

Next, we use the calculated energy of reaction, energy of synthesis, and the assumed energy-transfer efficiency to get the amount of energy equivalents of the electron donor that must be oxidized (A):

$$A = -\frac{\frac{\Delta G_p}{\varepsilon^n} + \frac{\Delta G_{pc}}{\varepsilon}}{\varepsilon \Delta G_r} = -\frac{37.2}{0.6(-63.4)} = 0.98 \quad (7)$$

This amount of the electron donor consumed for energy (A equivalents) was then used to estimate the maximum fraction of electron equivalents used for synthesis ($f_s^0$) and the minimum fraction of electron equivalents for energy production ($f_e^0$).

$$f_s^0 = \frac{1}{1+A} = \frac{1}{1+0.98} = 0.51 \quad (8)$$

$$f_e^0 = 1 - f_s^0 = 1 - 0.54 = 0.49 \quad (9)$$

Lastly, we created a balanced stoichiometry equation using the fractions for synthesis and energy production and the half reactions listed in Supplemental Material Equations 3-6 and then calculated the yield.

$$R = f_e(R_a - R_d^{energy}) + f_s(R_{cells} - R_c^{synthesis}) \tag{10}$$

$$H_2 + 0.26CH_3COOH + 0.12NH_4^+ + 0.10HCO^- + 0.15CO_2$$
$$\rightarrow 0.91H_2O + 0.25CH_4 + 0.10C_5H_7O_2N$$

$$Y = 0.10\, mol\ cells \left(113 \frac{g\ cells}{mol\ cells}\right) / \, 1\, mol\ H_2 = \ 11.57 \frac{g\ cells}{mol\ H_2} \tag{11}$$

For a more detailed explanation on how to calculate stoichiometry equations and yields using this standardized procedure, please refer to the Rittmann and McCarty Environmental biotechnology textbook (6).

It may be useful to note that the stoichiometry table in Supplemental Table 2 is in gCOD not mols, so the stoichiometry coefficients in Supplemental Equation 10 are multiplied by the specific chemical oxygen demand, or the *COD* (gCOD $mol^{-1}$) for each component.

### Supplemental Material References


1. Corbin KD, Carnero EA, Dirks B, Igudesman D, Yi F, Marcus A, Davis TL, Pratley RE, Rittmann BE, Krajmalnik-Brown R, et al. Host-diet-gut microbiome interactions influence human energy balance: a randomized clinical trial. Nature Communications 2023 14:1. Nature Publishing Group; 2023;14:1–17. Available from: https://www.nature.com/articles/s41467-023-38778-x
2. Dirks B, Davis TL, Carnero EA, Corbin KD, Smith SR, Rittmann BE, Krajmalnik-Brown R. Methanogens are associated with altered microbial production of short-chain fatty acids and human-host metabolizable energy. bioRxiv. Cold Spring Harbor Laboratory; 2025;2024.12.31.630929. Available from: https://www.biorxiv.org/content/10.1101/2024.12.31.630929v1
3. Stephen AM, Haddad AC, Phillips SF. Passage of carbohydrate into the colon. Direct measurements in humans. Gastroenterology. Elsevier Masson SAS; 1983;85:589–95. Available from: http://dx.doi.org/10.1016/0016-5085(83)90012-4
4. Rowan AM, Moughan PJ, Wilson MN, Maher K, Tasman-Jones C. Comparison of the ileal and faecal digestibility of dietary amino acids in adult humans and evaluation of the pig as a model animal for digestion studies in man. British Journal of Nutrition. 1994;71:29–42.
5. Ndou SP, Kiarie E, Walsh MC, Ames N, M de Lange CF, Nyachoti CM. Interactive effects of dietary fibre and lipid types modulate gastrointestinal flows and apparent digestibility of fatty acids in growing pigs. British Journal of Nutrition. 2018;121:469–80. Available from: https://doi.org/10.1017/S0007114518003434
6. Rittmann BE, McCarty PL. Environmental biotechnology. Technology Guide: Principles and Applications. 2nd ed. McGraw-Hill Book Co; 2020.

7. Samuel BS, Hansen EE, Manchester JK, Coutinho PM, Henrissat B, Fulton R, Latreille P, Kim K, Wilson RK, Gordon JI. Genomic and metabolic adaptations of Methanobrevibacter smithii to the human gut. Proc Natl Acad Sci U S A. Proc Natl Acad Sci U S A; 2007;104:10643–8. Available from: https://pubmed.ncbi.nlm.nih.gov/17563350/
